# REACT (Real-world Emulated Clinical Trials): A Framework for Drug Repurposing Applied to Osteoporotic Fractures

**DOI:** 10.64898/2026.03.20.26348832

**Authors:** Jianing Liu, Seungyeon Lee, Xianhui Chen, Soledad A. Fernández, Qing Wu, Ping Zhang

## Abstract

Osteoporotic fractures remain common despite available therapies, highlighting the need for new, safe, and affordable treatment options. Advances in real-world data and causal inference have enabled target trial emulation, with prior frameworks demonstrating drug repurposing using observational data. However, no large-scale trial emulation framework has been applied to systematic drug repurposing for osteoporotic fracture prevention. To address this gap, we developed REACT (Real-world Emulated Clinical Trials), a scalable framework that operationalizes established target trial principles into an automated, comparator-robust pipeline for high-throughput drug screening. Applied to fracture prevention in older adults with osteoporosis, REACT screened 1,222 drugs and identified 18 with statistically significant associations, including 11 with protective effects and 7 with risk-increasing effects. The convergence of these findings with biological and prior observational evidence underscores REACT’s ability to surface clinically meaningful patterns and its potential as a generalizable platform for accelerating real-world drug discovery and safety surveillance.

## Introduction

Osteoporosis is a chronic skeletal disorder characterized by low bone mineral density and microarchitectural deterioration of bone tissue, leading to an increased susceptibility to fractures—particularly of the hip, spine, and wrist. Osteoporotic fractures are a major public health burden, affecting approximately 1 in 2 women and 1 in 4 men over the age of 50 in the United States^1^. These fractures contribute significantly to loss of mobility, reduced quality of life, and increased mortality, especially in the elderly^2,3^.

Despite the availability of antiresorptive and anabolic therapies, current pharmacologic treatments have several limitations. Many patients experience side effects or fail to adhere to treatment regimens, and some therapies carry risks such as osteonecrosis of the jaw or atypical femur fractures^4^. Furthermore, a significant challenge is the limited number of novel drugs in the osteoporosis market, which results in a lack of treatment options for patients who do not respond to existing first-line agents^5^. As a result, treatment gaps persist, and fracture incidence remains high among older adults. In this context, drug repurposing – the systematic identification of new indications for existing, approved medications – offers a promising and cost-effective strategy to expand the therapeutic toolkit for osteoporosis^6^.

Recent advances in real-world data (RWD) and causal inference methods have made it possible to emulate randomized clinical trials (RCTs) using observational data^7,8^. Foundational work on target trial emulation has emphasized the explicit specification of eligibility criteria, treatment strategies, index date, and causal contrasts to reduce bias in observational analyses^9,10^. Building on these principles, several large-scale frameworks have demonstrated the feasibility of systematically emulating multiple trials using healthcare databases for drug evaluation and repurposing^11,12^. These approaches integrate new-user active-comparator designs and confounding adjustment techniques to approximate randomized comparisons at scale. To date, however, no large-scale target trial emulation framework has been applied to systematic drug repurposing for osteoporotic fracture prevention.

To address this gap, we developed REACT (Real-world Emulated Clinical Trials), a scalable trial emulation pipeline for high-throughput drug repurposing. REACT operationalizes established design and adjustment principles into an automated, comparator-robust pipeline that enables high-throughput screening of marketed medications within a unified fracture-focused analytic framework.

## Material and Methods

### Data Source

We used the MarketScan Medicare Supplemental and Coordination of Benefits (MDCR) Database (2016–2022)^13^, a large-scale, real-world longitudinal healthcare claims warehouse representing more than 8 million U.S. retirees aged 65 years or older. The database contains de-identified, patient-level information on diagnoses, procedures, prescription dispensations, and demographic characteristics.

From this dataset, we identified approximately 547,000 patients with osteoporosis using diagnosis records encoded in the International Classification of Diseases, Tenth Revision, Clinical Modification^14^ (ICD-10-CM). These patients carry a wide range of diagnosis codes reflecting their full clinical history. To organize this high-dimensional diagnostic information, all ICD-10-CM codes were mapped to the Clinical Classifications Software Refined (CCSR) system, a modern grouping methodology developed by the Agency for Healthcare Research and Quality. CCSR consolidates more than 70,000 ICD-10-CM diagnosis codes into 552 clinically meaningful categories. Drug exposures were captured through national drug codes (NDCs) and linked to ingredient-level identifiers.

### Study Cohort

A new-user design was adopted to ensure that exposure was clearly established and temporally aligned with covariate assessment and outcome follow-up, thereby reducing immortal time bias and misclassification of long-term or intermittent users^7,15^. Eligible patients were adults aged ≥65 years with a documented osteoporosis diagnosis^16,17^ (ICD-10 M81.x) and at least one year of continuous enrollment before cohort entry. Users were required to have ≥2 prescriptions for drug *d* separated by ≥30 days to ensure adequate exposure.

To estimate drug effects, we constructed parallel new-user active-comparator cohorts (Figure 1). For each target drug *d*, new users were compared with new users of randomly selected active medications meeting the same baseline eligibility criteria. The index date 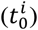 was defined as the first prescription following a 12-month washout period, and follow-up began at 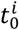 with person-time accrued thereafter. Both groups were required to have no prior osteoporotic fractures and at least two years of continuous follow-up for outcome assessment. Fractures were identified using ICD-10-CM codes corresponding to osteoporotic fracture sites as defined in Lippuner et al.^16^, with the first qualifying diagnosis during follow-up treated as the incident event.

**Figure 1.**
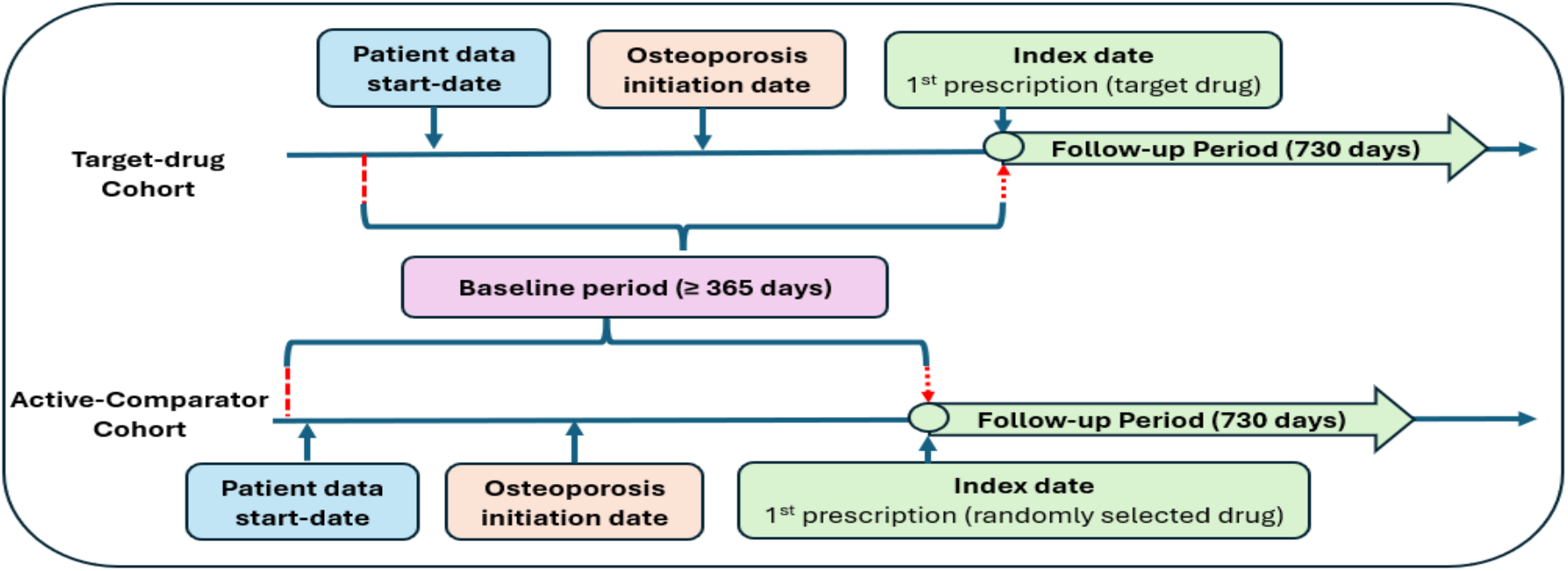
Cohort design timeline using Merative MarketScan (2016–2022)).

This design established a 12-month baseline period 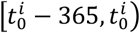 for covariate measurement and a two-year follow-up period 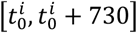 for outcome evaluation. Figure 1 illustrates the cohort structure and timeline.

### Study Design: The REACT Framework

We used the REACT framework to efficiently screen and emulate high-throughput RCT for candidate drugs. Figure 1 and Figure 2 together illustrates the end-to-end drug repurposing pipeline implemented in this study. Starting from EHR/claims data, REACT first applies predefined study design rules—including treatment assignment, eligibility criteria, adherence specifications, and outcome definitions—to automatically construct target-drug and active-comparator cohorts for each candidate drug. These rich longitudinal diagnosis, procedure, and medication histories are then converted into covariates that capture patients’ baseline clinical profiles. For every target drug, REACT executes an emulated RCT using IPTW to estimate causal treatment effects while adjusting for confounding.

**Figure 2.**
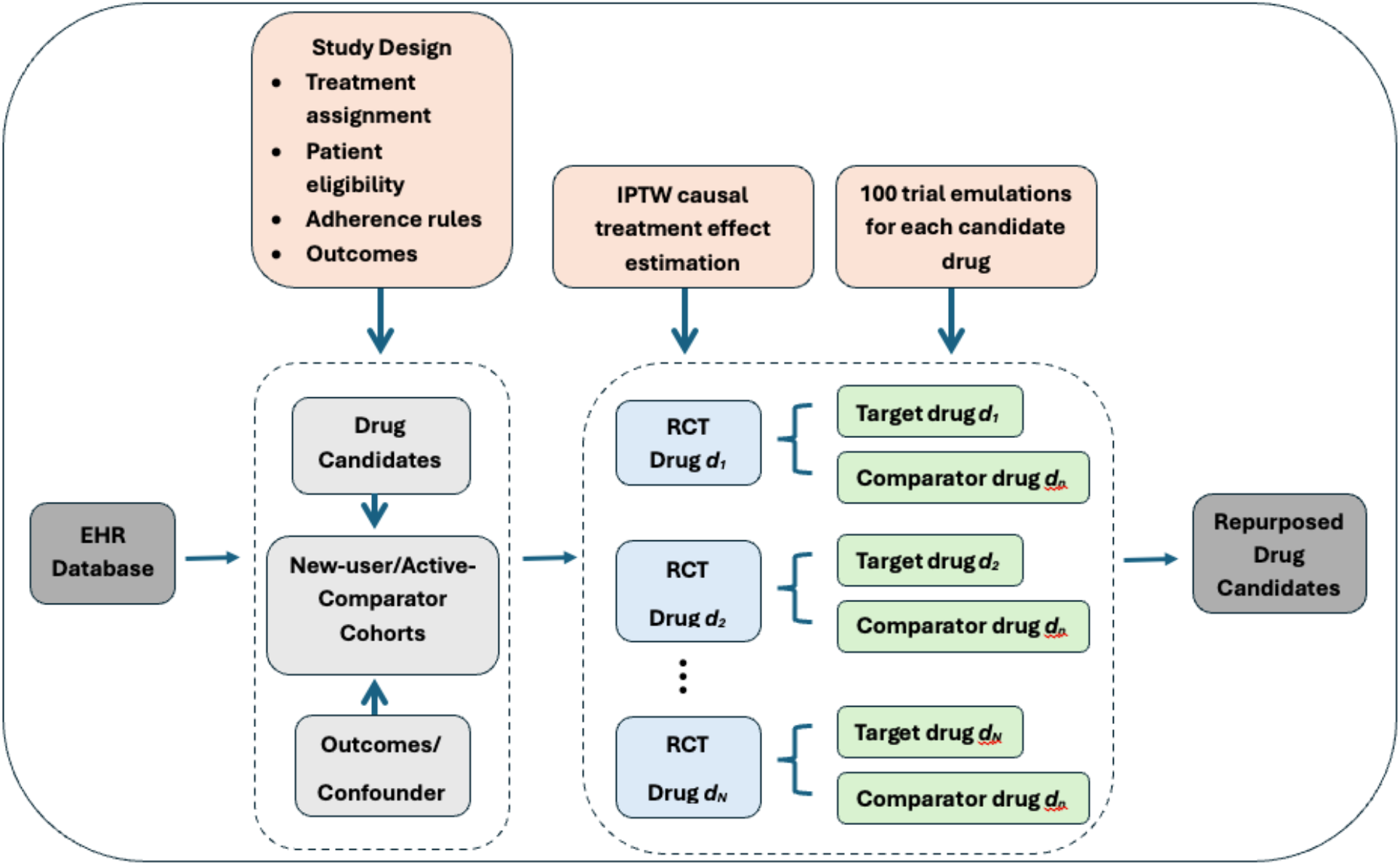
Overview of the high-throughput drug repurposing pipeline.

#### Treatment Assignment

For each candidate drug *d*, we emulated a two-arm trial comparing new users of target drug *d* with new users of an active comparator. Treatment assignment was defined using observed prescription claims. Patients were classified as:

- A_i_ = 1 (treated): individuals who initiated drug *d*_*1*_, defined as the first prescription date after a 12-month washout with no prior use of *d*_*1*_;
- A_i_ = 0 (control): individuals who initiated a different on-market drug *d*_*n*_ not indicated for osteoporosis and who never received *d*_*n*_ during the study period.

This design follows a new-user active-comparator approach, ensuring temporal alignment of treatment initiation and minimizing immortal time and selection biases.

#### Covariates and Balance Assessment

To construct diagnosis-based covariates, we mapped all ICD-10-CM codes in the baseline period to the 552 CCSR categories, which serve as the high-dimensional diagnosis feature set for confounding adjustment. Using CCSR allows us to retain important clinical detail while mitigating extreme sparsity and dimensionality inherent to raw ICD-10-CM codes, thereby improving model stability and interpretability during inverse probability weighting and covariate balance diagnostics^12^. Each CCSR category was modeled as a binary indicator variable (0/1) representing whether the patient had at least one diagnosis within that category during the baseline window. Specifically, for each patient i and CCSR category k, the covariate *X*_*ik*_ was defined as:

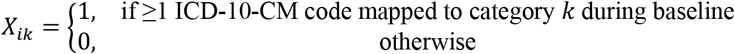

We constructed a high-dimensional covariate matrix *X*_*i*_ for each patient and estimated the propensity score *e*(*X*_*i*_) = *P*(*A*_*i*_ = 1|*X*_*i*_) using logistic regression. To adjust for baseline confounding, we applied inverse probability of treatment weighting^18^ (IPTW) with stabilized weights:

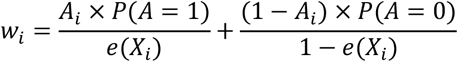

Weights were trimmed at the 1st and 99th percentiles to reduce the influence of extreme values^9^.

Covariate balance between treatment and control cohorts was assessed using the absolute standardized mean difference^19^ (SMD) for each covariate:

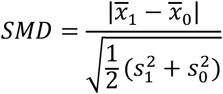

where 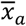 and 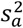 denote the weighted mean and variance of the covariate in group *A*_*i*_ = *a*. A trial was considered balanced if:

- All individual SMDs were ≤ 0.2^20^, and
- No more than 2% of covariates exceeded the threshold.

#### Effect Estimation and Statistical Significance

For each emulated trial, the average treatment effect^10^ (ATE) was estimated as the risk difference in the incidence of osteoporotic fracture between the weighted treatment and control groups:

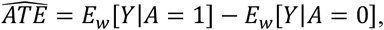

where E_*w*_ represents the weighted mean under IPTW.

For each drug, 100 independent target trial emulations were conducted using distinct randomly selected active comparators. The reported mean effect represents the average ATE across emulations. Statistical significance of the mean ATE was assessed using a one-sample t-test testing whether the average effect differed from zero. Confidence intervals were constructed using the empirical standard deviation of replicate ATE estimates, reflecting variability across emulated trial specifications. This repeated-emulation strategy captures uncertainty arising from alternative comparator selection and differences in covariate overlap and balance, thereby reducing dependence on any single comparator choice. The resulting distribution of ATE estimates reflects design-level heterogeneity rather than patient-level sampling variability.

#### Multiple Testing Correction

Because the screening pipeline evaluates a large number of candidate drugs, we applied a false discovery rate^21^ (FDR) correction to control for multiplicity. Raw p-values from the one-sample t-tests were adjusted using the Benjamini– Hochberg procedure. Drug–outcome associations with an FDR-adjusted p-value < 0.05 were classified as statistically significant and retained for further consideration.

#### Drug Selection Criteria

We performed large-scale, real-world screening for potential osteoporosis therapies using a high-throughput target trial emulation framework. Beginning with the complete set of candidate drugs, REACT sequentially applied eligibility filters as summarized in Figure 3. Each drug underwent comprehensive covariate balance diagnostics, evaluated using SMD before and after IPTW. Only drugs that achieved adequate post-weighting balance—indicating successful control of measured confounding—advanced to the effectiveness evaluation stage. Among these, drugs exhibiting a statistically significant ATE (FDR-adjusted p < 0.05) were designated as repurposing candidates, representing treatments with consistent, reproducible, and sufficiently precise evidence of impact on osteoporotic fracture risk.

**Figure 3.**
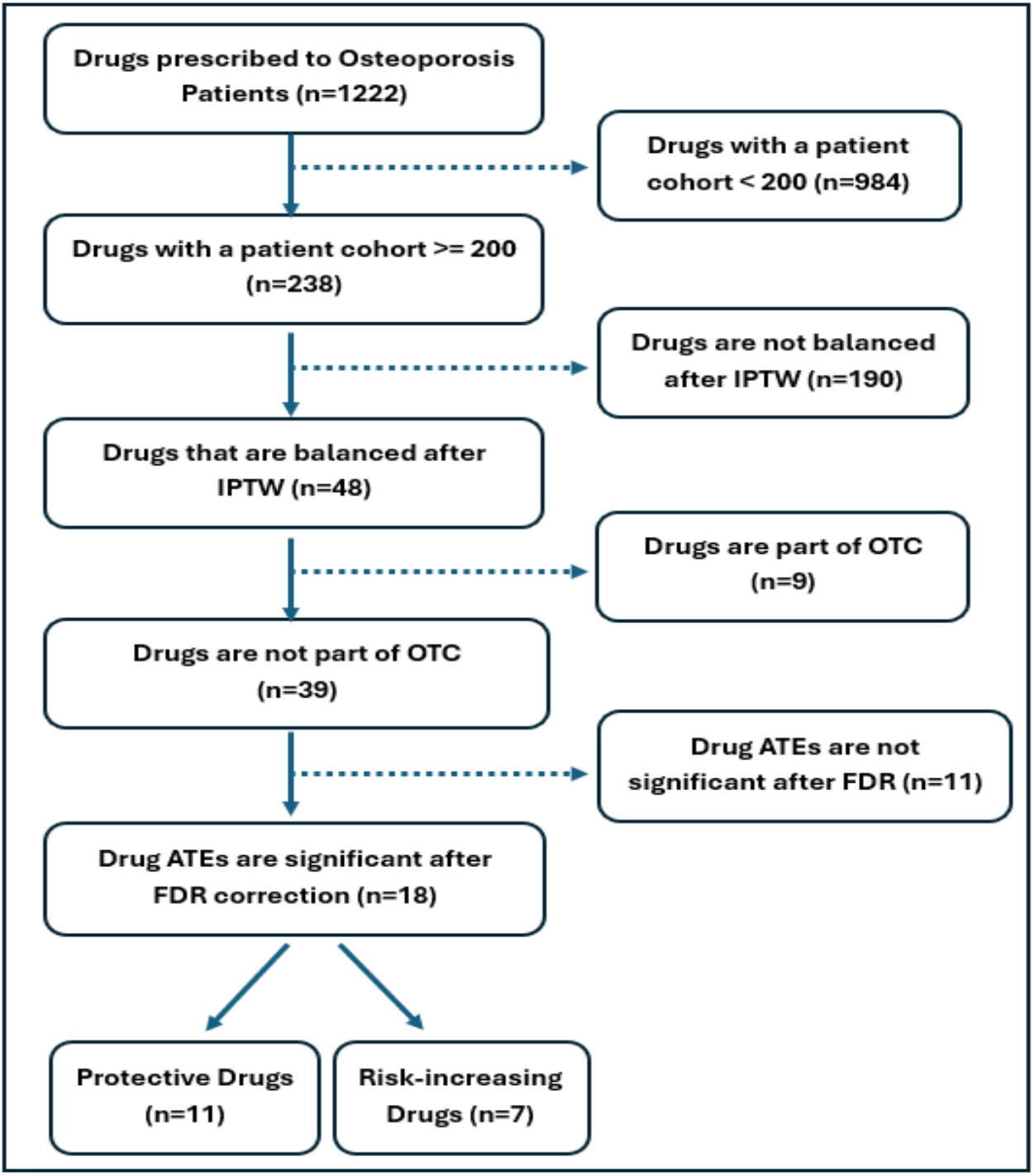
Drug selection and screening criteria. From 1,222 drugs in the dataset, REACT filters down to 18 drugs.

## RESULTS

We identified 1,222 drugs prescribed to patients with osteoporosis, of which 238 candidate drugs had sufficiently large cohorts (≥200 patients) to support emulated randomized trial analyses. After estimating treatment effects using IPTW, we evaluated covariate balance for each emulated trial. Forty-eight drugs met the prespecified balance criteria (<2% of covariates with SMD >0.20) and were carried forward for further evaluation. We then excluded medications primarily available over-the-counter, as their limited capture in claims data could lead to exposure misclassification and unreliable effect estimation. Among the remaining 39, 18 drugs remained significant after applying multiple-testing correction (FDR <0.05). Eleven drugs demonstrated protective effects with negative ATEs, and seven exhibited positive ATEs consistent with increased fracture risk.

### Protective Drug Candidates

The protective group included several medications that demonstrated statistically significant reductions in fracture risk, including Losartan (ATE = –0.024), Valsartan (ATE = –0.024), Pravastatin (ATE = –0.024), Rosuvastatin (ATE = –0.022), Montelukast, Amoxicillin, Spironolactone, Cefpodoxime, Irbesartan, Simvastatin, and Mupirocin. Although ARBs (e.g., Losartan, Valsartan, Irbesartan) and statins (e.g., Pravastatin, Rosuvastatin, Simvastatin) have accumulating observational and mechanistic evidence suggesting beneficial effects on bone remodeling, inflammation, and fracture risk, none of these agents are currently clinically indicated for osteoporosis or bone protection. Our findings therefore reinforce emerging but off-label therapeutic signals.

Consistent with prior work, we found evidence supporting the potential bone-protective roles of the following agents: (1) Losartan and other ARBs have been associated with reduced bone resorption and improved bone microarchitecture in both animal models and human observational studies^22,23^; (2) Valsartan has been linked to anti-inflammatory and anti-oxidative pathways that may attenuate osteoporosis progression^24^; (3) Pravastatin stimulates bone morphogenetic protein-2 (BMP-2) expression and has been linked to reduced fracture incidence in population studies^25^; (4) Rosuvastatin and other statins has demonstrated favorable effects on bone turnover markers and fracture risk in observational datasets^26,27^; (5) Montelukast exhibits anti-inflammatory effects that may indirectly support bone preservation^28^; (6) Spironolactone has been linked to markedly lower osteoporosis and fracture risk in hypertensive patients^29^. Collectively, these protective signals highlight several widely used, well-tolerated agents with plausible biological mechanisms or supportive epidemiologic evidence. Each merits further mechanistic investigation, validation in external datasets, and potential prioritization for prospective clinical evaluation.

**Table 1.** Protective Drug Candidates.

| Drug Name | Estimated Effect (Mean[CI]) | FDR p-value | No. of unbalanced features |
| --- | --- | --- | --- |
| Losartan | -0.024 (-0.028 - -0.019) | <0.0001 | 3.76 |
| Valsartan | -0.024 (-0.030 - -0.017) | <0.0001 | 4.59 |
| Pravastatin | -0.024 (-0.028 - -0.019) | <0.0001 | 2.79 |
| Rosuvastatin | -0.022 (-0.027 - -0.015) | <0.0001 | 5.4 |
| Montelukast | -0.019 (-0.025 - -0.014) | <0.0001 | 4.7 |
| Amoxicillin | -0.015 (-0.020 - -0.009) | <0.0001 | 3.08 |
| Spironolactone | -0.013 (-0.019 - -0.006) | 0.004 | 6.65 |
| Cefpodoxime | -0.012 (-0.017 - -0.006) | 0.003 | 5.54 |
| Irbesartan | -0.011 (-0.016 - -0.005) | 0.003 | 4.37 |
| Simvastatin | -0.008 (-0.012 - -0.003) | 0.003 | 2.82 |
| Mupirocin | -0.007 (-0.012 - -0.002) | 0.009 | 3.41 |

### Risk-Enhancing Drug Candidates

Conversely, seven drugs demonstrated significantly positive ATEs, indicating a potential increase in fracture risk among users. The risk-enhancing group included Tramadol, Alprazolam, Cyclobenzaprine, Salmeterol, Ipratropium, Levofloxacin, and Nitroglycerin. Several of these findings are consistent with prior safety literature. Tramadol has been associated with an increased risk of hip fractures in older adults, likely mediated by sedation and falls^30^. Benzodiazepines, including Alprazolam, are well known to increase fall-related injuries and hip fractures in older populations^31^, and skeletal muscle relaxants such as Cyclobenzaprine have similarly been linked to higher rates of fall and fracture events^32^. While confounding by indication cannot be ruled out for any of these agents, the rigorous covariate adjustment and balance diagnostics in our framework support these drugs as plausible safety signals and/or markers of elevated underlying fracture risk.

**Table 2.**
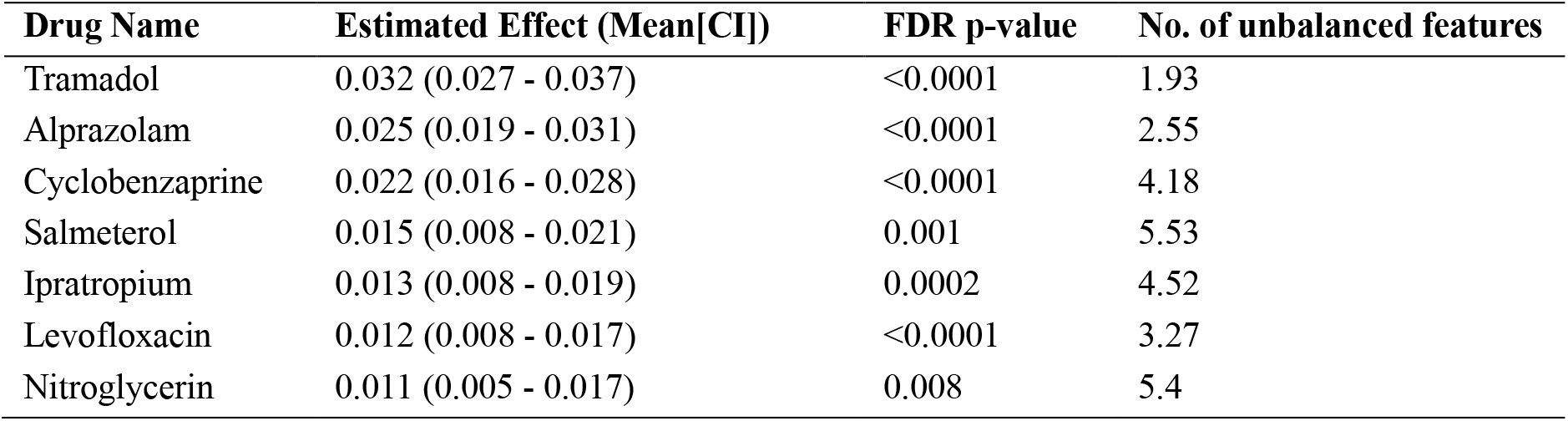
Risk-Enhancing Drug Candidates.

### Drug Effect Estimates Overview

To evaluate the magnitude and statistical significance of treatment effects across all 48 balanced drugs, we first generated a volcano plot (Figure 4), which summarizes both effect size and confidence in the estimated associations. The x-axis displays the Average Treatment Effect (ATE), where negative values represent a protective effect against osteoporotic fracture and positive values represent increased fracture risk. The y-axis shows the –log10(FDR-adjusted p-values), providing a direct visual indication of statistical strength. Drugs with strong, statistically significant protective effects appear in the upper-left quadrant in blue, while risk-increasing drugs appear in the upper-right quadrant in red. This visualization distinguishes protective vs. harmful signals and highlights where meaningful associations rise above background noise.

**Figure 4.**
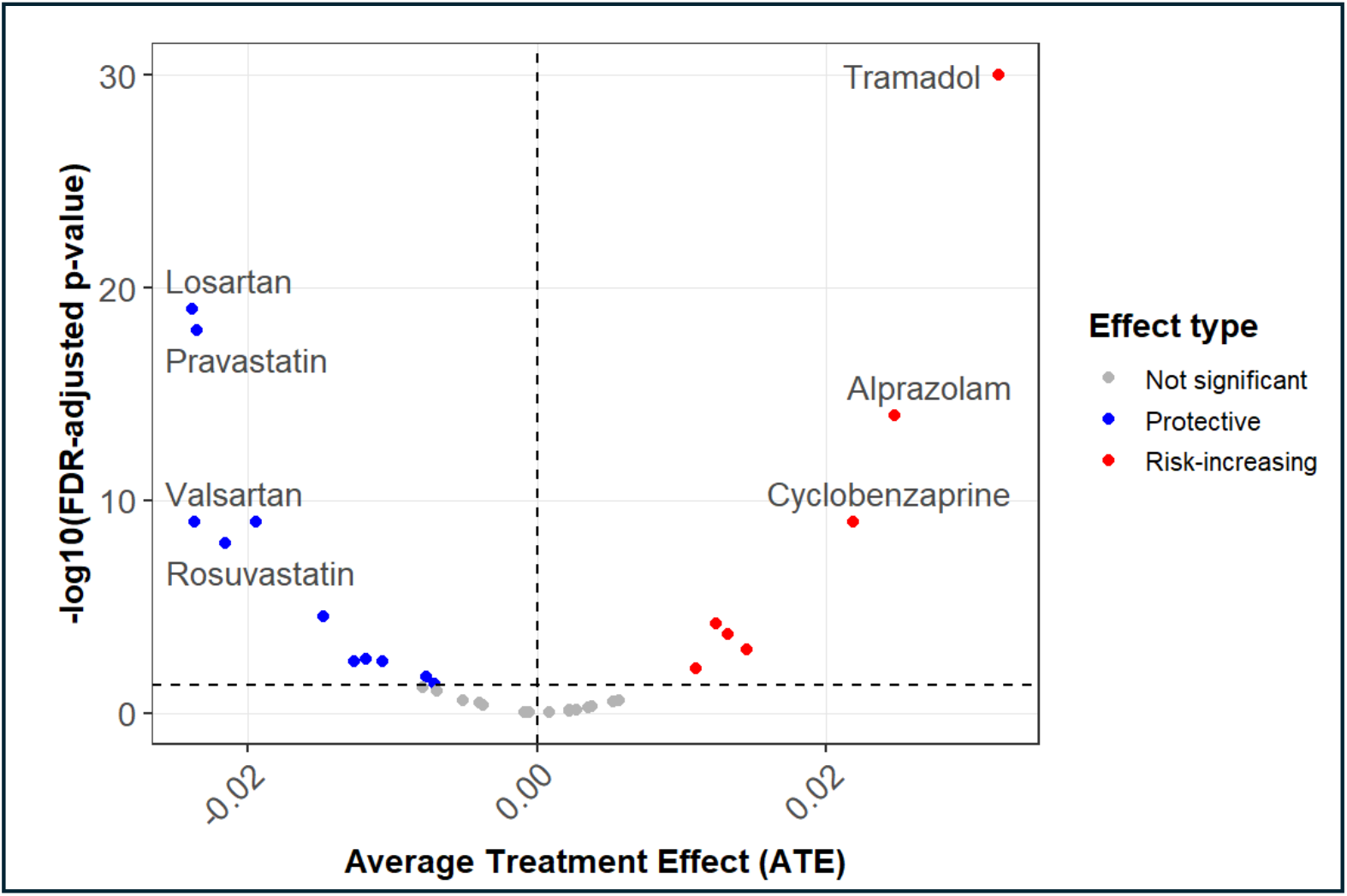
Volcano plot of drug effects.

Building on these results, we further examined the robustness and precision of treatment effect estimates for the 18 top candidates that passed screening criteria: (1) FDR-adjusted p ≤ 0.05, (2) all individual SMDs were ≤ 0.2, and (3) post-IPTW imbalance ≤ 2%. For these drugs, we constructed a forest-style boxplot summary (Figure 5) that displays the distribution of ATE estimates across 100 independent trial emulations using different active comparators. Each boxplot depicts the median, interquartile range, and variability of the ATE estimates across the 100 emulated trials for each significant drug, enabling direct comparison of effect magnitude and uncertainty among the final candidates. By summarizing results across multiple comparator-based emulations, the visualization reflects the stability of each drug’s estimated effect rather than relying on any single trial emulation.

**Figure 5.**
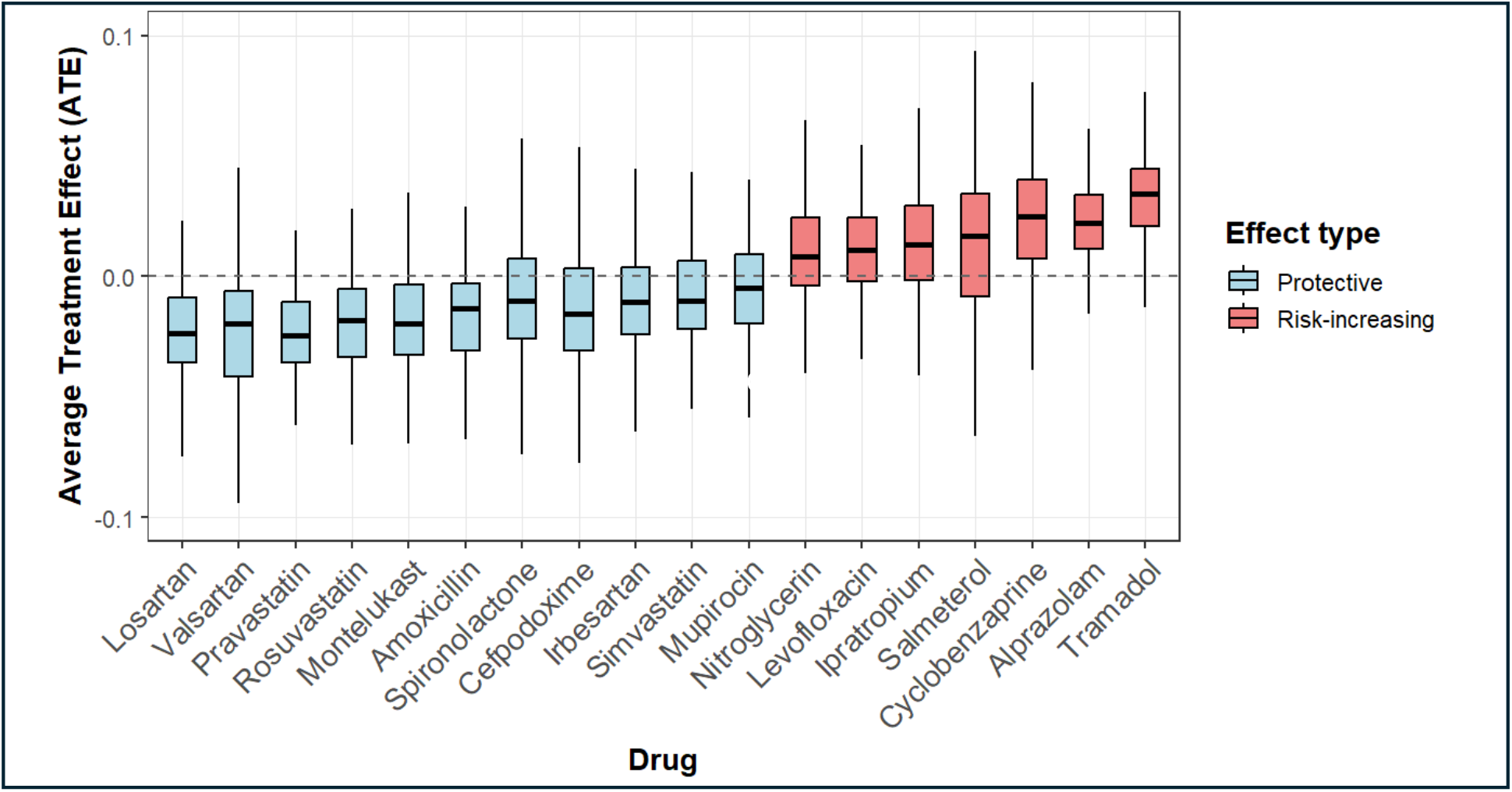
Distribution of estimated ATE of drugs on defined outcomes across 100 independent trial emulations.

Together, the volcano plot and the emulation-based boxplots provide a coherent picture of repurposing potential within the drug landscape. Protective agents—with consistently negative ATEs and supportive statistical evidence—cluster largely among cardiovascular medications such ARBs and statins. Conversely, medications with risk-increasing associations, including certain analgesics and muscle relaxants, display positive ATEs. This integrated view demonstrates that large-scale target trial emulation can reliably identify both promising repurposing candidates and clinically relevant safety signals for osteoporotic fracture prevention.

### Covariate Balancing

To illustrate the effectiveness of IPTW in achieving covariate balance, we examined *Valsartan* (one drug candidate from of the protective list) as a representative example. Figure 6 shows the absolute SMDs for the 20 most imbalanced baseline covariates before and after weighting. Prior to IPTW, several covariates displayed substantial imbalance (SMD >0.20), with some exceeding 0.40, reflecting marked differences in underlying clinical complexity between *Valsartan* new users and comparators. After applying IPTW, the absolute SMDs for nearly all covariates were substantially reduced, with the vast majority falling below 0.10. This example demonstrates how IPTW effectively aligns high-dimensional diagnosis-based covariates between treatment groups, thereby reducing confounding and strengthening the validity of the causal effect estimates in the *Valsartan* emulated trial.

**Figure 6.**
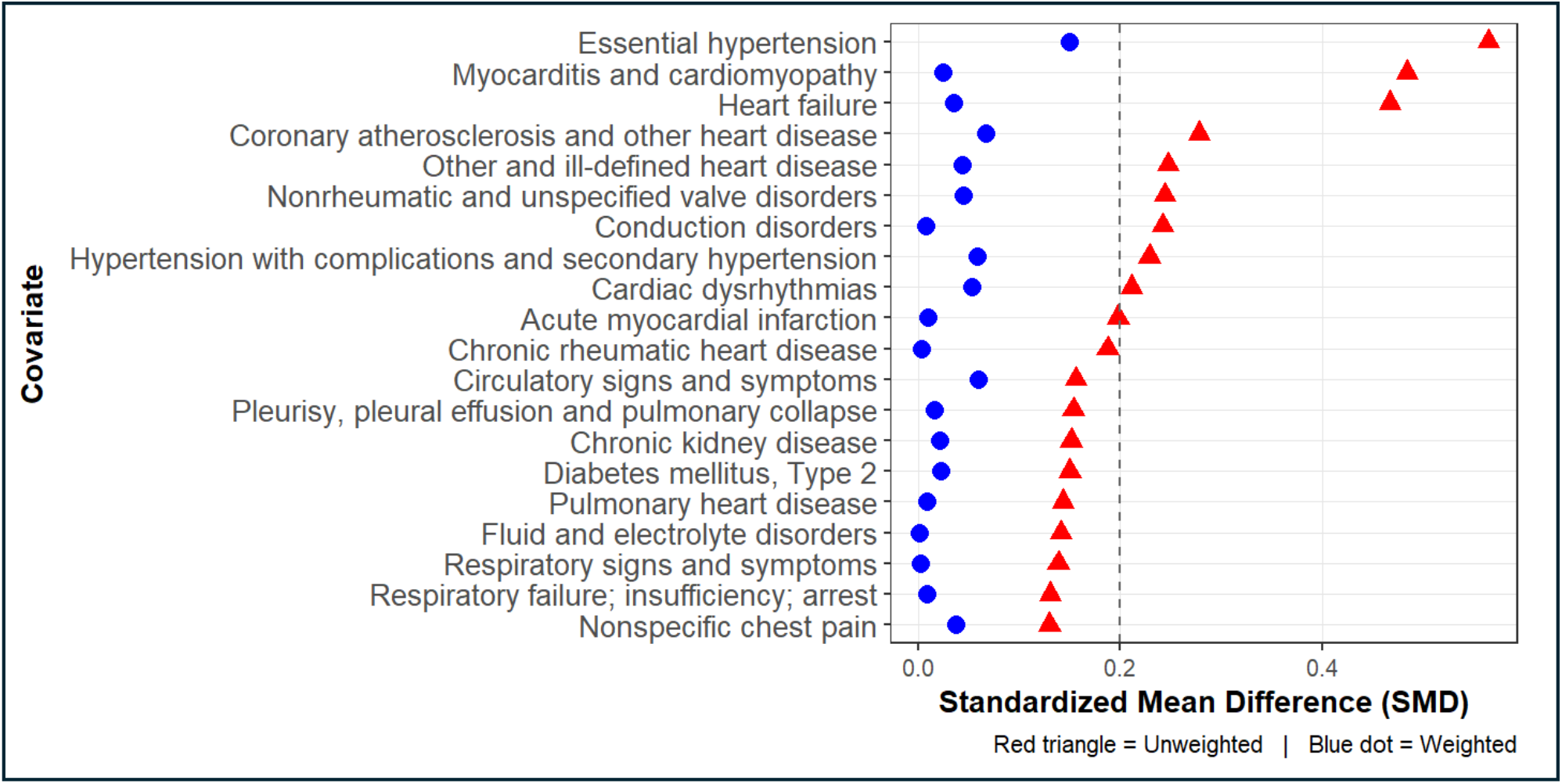
Top 20 Imbalanced Covariates Before and After IPTW for Valsartan (Drug 1308842).

## Conclusion

In this study, we applied the large-scale REACT trial-emulation framework to identify both repurposing candidates and potential safety signals for osteoporotic fracture prevention using real-world data. The recovery of mechanistically coherent protective drug classes—supported by established biological pathways^24,33^ and prior epidemiologic evidence for beneficial effects on bone health^23,34^—provides strong external validation of the framework’s ability to surface clinically meaningful, literature-consistent signals rather than spurious associations. Likewise, the identification of medications associated with increased fracture risk reflects interpretable pathways related to falls and impaired mobility^26,30^, further underscoring the robustness of the approach. At the same time, these findings should be interpreted as exploratory signals rather than definitive clinical recommendations. The identified candidates represent hypothesis-generating associations that require further validation in independent datasets and prospective study designs. In addition, comprehensive assessment of adverse events and overall risk–benefit profiles will be essential before considering clinical translation.

This work has limitations. Claims data lack granular clinical measures such as bone mineral density, frailty assessments, and DXA imaging, and exposure misclassification remains possible. Although IPTW reduces confounding, unmeasured bias may persist, and generalizability may be limited to commercially insured U.S. adults aged ≥65 years. Future research may incorporate formal negative control outcome analyses and quantitative sensitivity analyses for unmeasured confounding to further strengthen causal interpretation. Additional methodological extensions may include cumulative exposure modeling, duration-stratified analyses, time-varying exposures, integration of EHR-derived phenotypes, and external validation through prospective or pragmatic trial designs. Despite these constraints, the strong convergence between REACT’s top signals and existing biological and epidemiologic evidence provides external validation of the framework’s ability to surface meaningful associations at scale. Overall, REACT offers a practical and rigorous approach for uncovering repurposing opportunities and safety concerns across therapeutic domains, underscoring how structured causal inference pipelines can accelerate evidence generation in real-world settings.

## Data Availability

The data used in this study are from Merative MarketScan and were obtained under a data use agreement. These data are not publicly available and cannot be shared by the authors. Researchers may obtain access to the data directly from Merative MarketScan with appropriate permissions.

## Acknowledgement

This study was supported by the National Science Foundation (IIS-2145625) and the National Institute on Aging (R01AG080017).

